# In vivo Assessment of Neuroinflammation in 4-Repeat Tauopathies

**DOI:** 10.1101/2020.07.14.20150243

**Authors:** Carla Palleis, Julia Sauerbeck, Leonie Beyer, Stefanie Harris, Julia Schmitt, Estrella Morenas-Rodriguez, Anika Finze, Alexander Nitschmann, Francois Ruch-Rubinstein, Florian Eckenweber, Gloria Biechele, Tanja Blume, Yuan Shi, Endy Weidinger, Catharina Prix, Kai Bötzel, Adrian Danek, Boris-Stephan Rauchmann, Sophia Stöcklein, Marcus Unterrainer, Nathalie L. Albert, Christian Wetzel, Rainer Rupprecht, Axel Rominger, Peter Bartenstein, Jochen Herms, Robert Perneczky, Christian Haass, Johannes Levin, Günter U. Höglinger, Matthias Brendel

## Abstract

**Objective:** The aim of this cross-sectional single center study was to investigate 18kDa translocator protein (TSPO)-PET as a biomarker for microglial activation in the 4-repeat tauopathies corticobasal degeneration and progressive supranuclear palsy (PSP).

**Methods:** Specific binding of the TSPO tracer ^18^F-GE-180 was determined by serial PET during pharmacological depletion of microglia in a 4-repeat tau mouse model. TSPO-PET was performed in 30 patients with corticobasal syndrome (CBS, 68±9 years, 16 female) and 14 patients with PSP (69±9 years, 8 female), and 13 control subjects (70±7 years, 7 female). Group comparisons and associations with parameters of disease progression and sTREM2 were assessed by region-based and voxel-wise analyses.

**Results:** Tracer binding was significantly reduced after pharmacological depletion of microglia in 4-repeat tau mice. Elevated TSPO labeling (standardized-uptake-value-ratios) was observed in subcortical brain areas of CBS and PSP patients when compared to controls, most pronounced in the globus pallidus internus (CBS: 1.039 [95%CI 1.000–1.078, p<0.001], PSP: 1.046 [95%CI: 0.990–1.101, p<0.001], controls: 0.861 [95%CI 0.802–0.921]), whereas only CBS patients showed additionally elevated tracer binding in motor and supplemental motor areas. TSPO labeling was only correlated weakly with parameters of disease progression in CBS and PSP but allowed sensitive detection of 4-repeat tauopathy patients. sTREM2 did not differ between patients with CBS and controls.

**Interpretation:** Our data indicate a potential of ^18^F-GE-180 PET to detect microglial activation in the brain of 4-repeat tauopathy patients, fitting to predilection sites of the phenotype. TSPO-PET may serve as a sensitive early disease stage biomarker in 4-repeat tauopathies.

## Introduction

Four-repeat (4R) tauopathies encompass a variety of neurodegenerative diseases^1^, including the movement disorders corticobasal syndrome (CBS)^2^ and progressive supranuclear palsy (PSP)^3^. Both are devastating neurodegenerative disorders, leading to death on average within six to ten years after symptom onset^1^. The majority of CBS and PSP patients referred to autopsy are characterized by intracellular aggregates of 4R tau in neurons and astroglia. There are even suggestions to classify CBS and PSP as the same disease entity, termed 4R-tauopathy, with different topological manifestation in the brain^4^. However, there is substantial neuropathological overlap of the two syndromes and with Alzheimer’s disease and frontotemporal dementia^5^.

Neuroinflammation plays a major role in 4R tauopathies, although there are ongoing debates on cause and effect of glial contribution to disease^6^. Recovering reduced phagocytic activity of microglia late in the disease course reduces tau spreading in human brain tissue and mouse models^7, 8^, but microglia and the inflammasome contribute to tau spreading by increasing its propagation and aggregation^9, 10^. Thus, a better understanding of the role of neuroinflammation across different disease stages will be key for developing effective therapeutic strategies. Biomarkers allowing quantification of neuroinflammation in 4R tauopathies could ultimately unravel associations between neuroinflammation and clinical progression *in vivo*, resulting in a monitoring tool for immunomodulatory therapies.

Despite the incremental use of 18kDa translocator protein positron-emission-tomography (TSPO-PET) imaging in Alzheimer’s disease^11^ and β-amyloid mouse models^12, 13^, only two studies were performed so far in 4R tauopathies^14, 15^, both reporting higher ^11^C-PK11195 binding in subcortical target regions of PSP patients compared to controls. The detailed role of TSPO as a biomarker of activated microglia in relation to the different 4R tauopathy phenotypes and clinical progression has yet to be explored. Furthermore, ^11^C labelled radiotracers likely hamper the translation of study results into clinical practice or therapeutic trials due to the short half-life.

Thus, we aimed to investigate the potential of ^18^F-GE-180 TSPO-PET as a biomarker of microglial activation in 4R tauopathies. Pharmacological depletion of microglia was performed in a 4R tau mouse model to validate the specificity of the tracer. Dynamic scanning and non-invasive kinetic modelling in a subset of patients served to determine short acquisition windows for low patient burden and cost-effective TSPO-PET imaging. Finally, we recruited a substantial number of 4R tauopathy patients and controls to test the capability of ^18^F-GE-180 TSPO-PET to detect elevated microglia activation in CBS and PSP patients and to study associations of TSPO labeling with parameters of disease progression.

## Material and Methods

### TSPO-PET imaging before and after microglia depletion in a 4R tau mouse model

All animal experiments were performed in compliance with the National Guidelines for Animal Protection, Germany and with the approval of the regional animal committee (Regierung von Oberbayern) overseen by a veterinarian. Animals were housed in a temperature- and humidity-controlled environment with 12h light-dark cycle, with free access to food and water. Small animal PET (µPET) experiments were carried out in five female human tau P301S mice, a mouse line expressing the human 0N4R tau isoform with the P301S mutation in exon 10 of *MAPT* gene under control of the murine thy1 promoter^16^. TSPO-µPET examinations were performed at baseline (7 months of age) and during microglial depletion after PLX5622 administration for 7 weeks. The sample size calculation was based on in house TSPO-PET quantification in the same model with the baseline acting as control to archive a power (1–β) of 0.80 at α of 0.05. PLX5622 was provided by Plexxikon (Berkeley, CA, USA) and formulated in AIN-76A standard chow by Research Diets (New Brunswick, NJ, USA) at 1200 ppm. µPET imaging was performed as reported previously^17^. In brief, all mice were anesthetized with isoflurane (1.5%, delivered at 3.5 L/min) and were placed in the aperture of the Inveon PET system (Siemens, Erlangen, Germany). ^18^F-GE-180 TSPO-µPET with an emission window of 60-90 min p.i. was used to measure cerebral TSPO. All analyses were performed using PMOD V3.5 (PMOD technologies, Basel, Switzerland). After co-registration to an MRI mouse atlas as described before^18^, normalization of images to standardized-uptake-value (SUV) images was conducted. Predefined bilateral cortical (24 mm^3^), brainstem (12 mm^3^) and whole brain (500 mm^3^) target volumes of interest (VOIs) were used and SUV were compared between baseline and follow-up.

### Study design, study population and clinical assessments

Thirty patients with possible (n=12) or probable (n=18) CBS, 14 patients with possible (n=1) or probable (n=13) PSP Richardson syndrome according to Armstrong Clinical Research and Movement Disorders Society criteria respectively^2, 19^ and 13 β-amyloid negative control subjects without objectified cognitive impairment and with intact motor function were recruited at the University Hospital, LMU Munich between September 2017 and January 2020 (**Fig. 1**). Three 4R tauopathy (n=2 CBS, n=1 PSP) patients and one control were excluded prior to the analysis due to low affinity TSPO binding as revealed by genotyping of the rs6971 single nucleotide polymorphism (SNP). Nine CBS patients were excluded due to β-amyloid positivity in ^18^F-flutemetamol PET, and four PSP due to a non-Richardson syndrome/non-CBS phenotype. The study and the data analyses (ethics-applications: 17-569, 17-755 & 19-022) were approved by the local ethics committee (LMU-Munich, Germany). All participants provided written informed consent according to the Declaration of Helsinki. Clinical data were collected according to the German multi-center prospective ProPSP cohort study^20^. Disease duration was defined as the time between symptom onset and PET imaging. The PSP rating scale (PSPRS) served as disease severity parameter and the Montreal Cognitive Assessment (MoCA) was used to assess the severity of cognitive deficits. Schwab and England Activities of Daily Living (SEADL) were recorded as a global score of functional ability.

**Figure 1.**
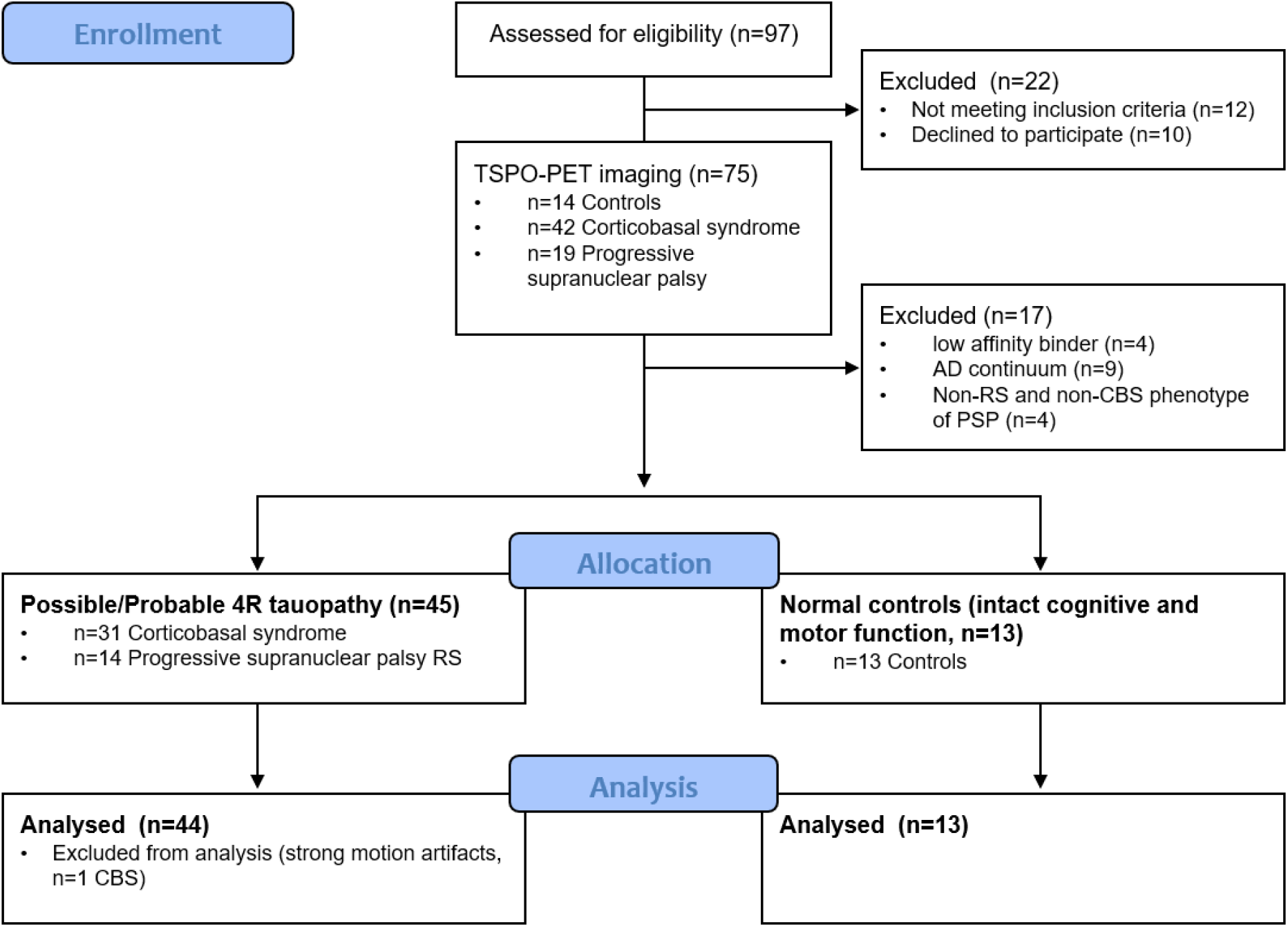
CONSORT chart of patient enrollment, allocation and analysis. CBS = corticobasal syndrome; PSP = progressive supranuclear palsy; RS = Richardson syndrome; AD = Alzheimer’s disease

### Human PET imaging

#### TSPO-PET acquisition

Patients were scanned at the Department of Nuclear Medicine, LMU, using a Biograph 64 PET/CT scanner (Siemens, Erlangen, Germany). Before the PET acquisition a low-dose CT scan was performed for attenuation correction. Emission data were either acquired dynamically over 90 min or statically from 60-80 min p.i. as described in detail below, starting with the injection of 189±12 MBq ^18^F-GE-180 as an intravenous bolus. The specific activity was 1714±523 GBq/μmol at the end of radiosynthesis, and the injected mass was 0.13±0.05 nmol. Images were reconstructed using a 3-dimensional ordered subsets expectation maximization algorithm (16 iterations, 4 subsets, 4 mm gaussian filter) with a matrix size of 336×336×109, and a voxel size of 1.018×1.018×2.027 mm. Standard corrections for attenuation, scatter, decay and random counts were applied. Dynamic PET data were systematically corrected frame-wise for subject motion within the PMOD Fusion tool (v3.5; PMOD Technologies, Zurich, Switzerland).

^18^F-GE-180 PET imaging was performed in a full dynamic setting (0–90 min p.i.) for a mixed population of eleven 4R tauopathy patients (6 CBS, 5 PSP) to allow non-invasive kinetic modelling and evaluation of a suitable time window for patient comfort and economic imaging. All other patients and controls received a static 60-80 min p.i. scan. The SUV 60-80 min p.i. images of all 44 4R tauopathy patients were averaged and compared to the average SUV of the 13 controls by calculating a percentage difference map. After visual and quantitative inspection (application of Hammers atlas regions), the bilateral antero-lateral temporal lobe was deemed suitable for further evaluation as a pseudo-reference tissue as it showed no differences between 4R tauopathy patients and controls and low variance. For dynamic datasets, the Logan reference tissue model in PMOD^21^ was used to calculate distribution volume ratio images (DVR; DVR = non displaceable binding potential (BPND) +1). As validated previously^22^, T* was set to 20 min, and the population average rate of k2’ was set to 0.027 1/min assuming similar average k2’ values for the antero-lateral temporal lobe when compared to the published values in the frontal cortex^23^. The antero-lateral temporal lobe also served as a reference tissue for calculation of SUV ratio (SUVr) images.

#### TSPO-PET data analysis

^18^F-GE-180 DVR and SUVr values were obtained in seven subcortical target regions^24^ defined by the atlas of the basal ganglia^25^, including an additional manually drawn midbrain target region^26^ and twelve cortical target regions as predefined by the Hammer’s atlas^27^ in the Montreal Neurology Institute (MNI) space. Subcortical target regions were defined as globus pallidus (internal & external part), putamen, subthalamic nucleus, substantia nigra, dentate nucleus^25, 27^, and midbrain. Cortical target regions were defined as precentral gyrus, superior frontal gyrus, middle frontal gyrus, inferior frontal gyrus, anterior orbital gyrus, medial orbital gyrus, lateral orbital gyrus, posterior orbital gyrus, straight frontal gyrus, anterior cingulate gyrus, postcentral gyrus and parietal cortex. Single frame SUVr values of dynamic datasets were correlated with 0-90 min DVR deriving from Logan graphical analysis. For target region-based group analyses the maximum ^18^F-GE-180 SUVr value of bilateral target regions was used to account for potential side asymmetries.

Statistical parametric mapping (SPM, V12, Functional Imaging Laboratory, The Wellcome Trust Centre in the Institute of Neurology, University College of London, UK) running in Matlab version R2016a (The MathWorks Inc., Natick, MA, USA) was used for voxel-wise statistical analyses. Group comparisons of ^18^F-GE-180 SUVr images of the full 4R tauopathy cohort as well as CBS and PSP patient groups versus controls were performed by an unpaired t-test using age, sex, and the TSPO polymorphism as covariates. Two levels of cluster significance were determined: A liberal SPM-threshold of p<0.05 without correction for multiple comparisons and a SPM-threshold of p<0.05 with strict family wise error correction for multiple comparisons.

### β-amyloid-PET acquisition and analysis

All CBS and control subjects underwent static ^18^F-flutemetamol PET imaging from 90 to 110 min p.i. as described above using the same PET scanner. Images were dichotomized into positive or negative by a visual read of a single nuclear medicine expert supported by regional Z-scores in frontal, parietal and temporal target regions as derived by the Gold software package (V4.17, HERMES Medial Solutions AB, Stockholm, Sweden).

### DNA extraction and SNP genotyping

Since the binding properties of second-generation TSPO ligands have been found to depend on genetic polymorphism of the TSPO gene^28, 29^, all individuals underwent rs6971 SNP genotyping and were classified as low, medium, or high affinity binder (LAB, MAB or HAB). For this purpose, whole-blood samples were sent to the Department of Psychiatry of the University Hospital Regensburg for genotyping. Genomic DNA was extracted from 4 ml of whole blood using a QIAamp DNA Blood Maxi kit (Qiagen, Hilden, Germany) according to the manufacturer’s protocol. DNA quality was assessed by optical absorbance and gel electrophoresis. Exon 4 of the TSPO gene and exon/intron junctions were amplified by PCR and sequenced using the Sanger method with the following primers: ex4-FAGTTGGGCAGTGGGACAG and ex4-R-CAGATCCTGCAGAGACGA. Sequencing data were analysed using SnapGene software (GSL Biotech; http://snapgene.com).

### sTREM2 measures

sTREM2 concentration was measured in the available cerebrospinal fluid samples by a modified assay based on the previously described sTREM2 ELISA using the MSD platform^30,31^. This assay was designed to selectively detect sTREM2 coming from cleavage of the full-length protein. In brief, we used Streptavidin-coated 96-well plates (MSD Streptavidin Gold Plates, cat. no. L15SA); a biotinylated polyclonal goat IgG anti-human TREM2 antibody (R&D Systems, cat. no. BAF1828; 0.25 µg/m) as capture antibody; a monoclonal rat IgG anti-human cleaved sTREM2 antibody (2 µg/mL) as a detection antibody, which is raised specifically against the cleavage site of human TREM2, recognizing the C-terminus sequence (EDAHVEH) from the cleaved fragment; and a SULFO-TAG-labeled goat polyclonal anti-rat IgG secondary antibody (MSD, cat. no. R32AH; 0.5 µg/mL). Recombinant TREM2 protein corresponding to the soluble cleaved fragment of human TREM2 (aminoacid 19-157) was purified from the supernatants of HEK293T cells stably overexpressing the protein and was used as standard. All antibodies, were diluted in the assay buffer (1% BSA and 0.05% Tween 20 in PBS, pH=7.4). The standard, the blanks, and the cerebrospinal fluid samples (duplicates; dilution, 1:6) were diluted in the same assay buffer supplemented with protease inhibitors (Sigma; Cat. # P8340). The plates were blocked overnight at 4°C in blocking buffer [3% bovine serum albumin (BSA) and 0.05% Tween 20 in PBS (pH 7.4); 200 μL/well] and next incubated with the capture antibody (25 µL/well) for 90 minutes at RT. After four washing steps with wash buffer (300 μL/well; 0.05% Tween 20 in PBS), the standard, the blanks, and the cerebrospinal fluid samples (50 μL/well) were incubated for 2 hours at RT. Plates were again washed six times followed by an incubation for 1 hour at RT with the detection antibody (50 µL/well). After six additional washing steps, plates were incubated with SULFO-tag conjugated secondary antibody (25 µL/well) for 1 hour in the dark at RT. Thereafter, plates were washed six times with wash buffer followed by two washing steps in PBS (300 μL/well). The light emission after adding 150 μL/well MSD Read buffer T (Cat. # R-92TC) was measured using the MESO QuickPlex SQ 120. All cerebrospinal fluid samples were distributed randomly across two plates and measured in the same day. Raw values are provided as ng/mL.

### Statistics

SPSS (V25, IBM, Ehningen, Germany) was used for statistical testing. ^18^F-GE-180 SUV of P301S mice were compared between baseline and microglial depletion by a paired Student’s *t*-test. In the subset of human dynamic scans, the agreement between DVR and SUVr of different time frames was assessed by Pearson’s coefficient of correlation (*R*). In the full human dataset, CoVs were calculated for late-phase ^18^F-GE-180 SUV and SUVr using the antero-lateral temporal reference tissue. Regional CoVs were compared between SUV and SUVr by a paired Student’s *t*-test. Age, PSPRS, disease duration, and MoCA scores were compared between the different study groups (CBS, PSP, controls) by a one-way analysis of variance, whereas sex and the rs6971 polymorphism were subject to a χ^2^ test. ^18^F-GE-180 SUVr of predefined target regions (maximum value of bilateral regions) were compared between the three study groups by a multivariate analysis of covariance including age, sex and rs6971 polymorphism as covariates as well as *post hoc* Bonferroni correction for multiple testing of groups. Raw sTREM2 measures were log transformed prior to comparison between patients with CBS and controls by a univariate analysis of covariance, including age and sex as covariates. A summed z-score vector was calculated for all 4R tauopathy patients by addition of single region z-scores ([individual SUVr patient – mean value SUVr controls] / standard deviation SUVr controls). For region-based classification, regional SUVr ≥ mean value +2 standard deviations of the controls were defined as positive. Here, one positive target region defined the subject as positive (dichotomous). Partial correlations were calculated for ^18^F-GE-180 SUVr in predefined regions and the summed z-score vector with clinical severity (PSPRS), disease duration and MoCA, controlled for age, sex and rs6971 polymorphism. Partial correlations were calculated for sTREM2 with clinical severity (PSPRS), disease duration and MoCA, controlled for age and sex. Flexible fitting models we calculated between the summed z-score vector and sTREM2 for patients with CBS. *P* values <0.05 were considered significant.

## Results

### Specificity of ^18^F-GE-180 to 4R tau associated microglia and implementation of an efficient TSPO-PET scan protocol

P301S 4R tau mice were imaged before and after microglial depletion by a CSF1R inhibitor. CSF1R blocking strongly reduced ^18^F-GE-180 binding in the whole brain of five P301S mice and model specific target regions (**Figs. 2A, B**), indicating specific detection of activated microglia by the tracer. Immunohistochemistry confirmed microglial depletion >90% by Iba1. Dynamic imaging in 4R tauopathy patients indicated a moderate brain uptake and fast wash out in 4R tauopathy target regions and the temporal reference region (**Fig. 2C**). SUVr tended to plateau after 60 minutes post injection (**Fig. 2D**), similar to earlier observations in rodents^32^. Agreement between Logan DVR from kinetic modelling and single frame SUVr reached *R*>0.6, starting >6 min post injection and dropped towards *R*<0.5 for frames >80 min post injection (**Fig. 2E**). 60-80 min SUVr overestimated 0-90 min DVR by 22.2±4.9% (cortical) and 29.5±11.0% (subcortical), but both correlated in a near linear fashion for cortical (*R*=0.91, *p*<0.001) and subcortical (*R*=0.85, *p*<0.001) target regions (**Fig. 2F**). Visual image analysis revealed highly similar uptake patterns of DVR and SUVr maps for cortical and subcortical TSPO labeling in 4R tauopathy patients (**Fig. 2G**). Percentage difference maps of SUV and SUVr indicated a high similarity of magnitude (**Fig. 1H**) but inter-subject variance was significantly decreased in the control and 4R tauopathy groups by the SUVr approach (**Fig. 1I**).

**Figure 2.**
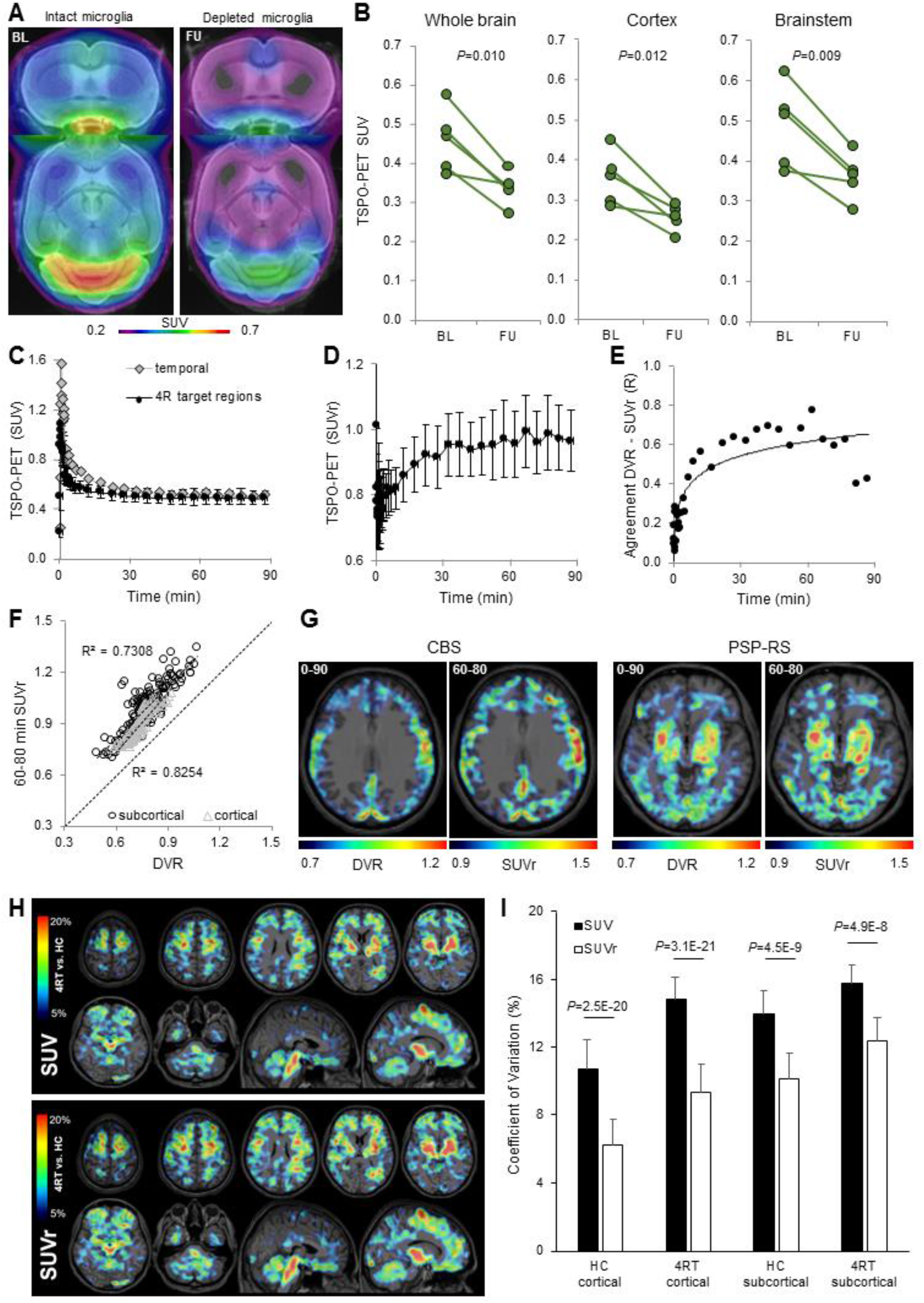
Preclinical validation of tracer specificity and methodological considerations for human 4R tauopathy TSPO-PET imaging: (**A**) Coronal and axial ^18^F-GE-180 TSPO-PET images of P301S mice upon an MRI standard template show strong decreases of the cerebral tracer uptake in week 7 of microglia depletion (FU) when compared to baseline (BL). Images represent the average standardized uptake value (SUV) of n=5 P301S mice. (**B**) Spaghetti plots show individual TSPO-PET SUV changes between baseline and a state of depleted microglia in target regions of P301S mice. *P*-values derive from a paired Student’s t-test. (**C**) Time-activity-curves of 4R tauopathy target regions (mean value ± standard deviation of 19 brain regions) and a temporal lateral pseudo-reference tissue. (**D**) Time-activity-ratio-curves of 4R tauopathy target regions divided by binding in the pseudo-reference tissue (mean value ± standard deviation of 19 brain regions). (**E**) Agreement between distribution volume ratios (DVR) and SUV ratios (SUVr) in single frames over the 90-minute scan duration. Each dot represents the mean Pearson’s correlation coefficient (R) of 19 4R tauopathy target regions. (**F**) Agreement of DVR with 60-80-minute SUVr of cortical and subcortical 4R tauopathy target regions as quantified at the single patient level. Values of **C-F** derive from n=11 4R tauopathy patients (n=6 corticobasal syndrome, n=5 progressive supranuclear palsy). (**G**) Individual examples of visual agreement between DVR and 60-80-minute SUVr for a patient with corticobasal syndrome (CBS, 58y, male, PSP Rating Scale: 27, disease duration: 37 months; MoCa: 14) and a patient with progressive supranuclear palsy Richardson syndrome (PSP-RS, 74y, male; PSP Rating scale: 18; disease duration: 10 months; MoCa: 22). Axial planes of ^18^F-GE-180 TSPO-PET images are projected upon a standard MRI atlas. (**H**) Percentage difference maps of n=44 4R tauopathy patients (4RT) and n=13 controls for SUV and SUVr quantification of the ^18^F-GE-180 TSPO-PET group average. Percentage differences are projected in axial and sagittal planes upon a standard MRI atlas. (**I**) Coefficients of variance (CoV) in the contrast of SUV and SUVr quantification. Bars represent mean CoV values ± standard deviation of cortical (n=12) and subcortical (n=7) 4R tauopathy target regions. *P*-values derive from a paired Student’s t-test.

### TSPO labeling follows the expected topology in CBS and PSP patients

All demographics and disease parameter are provided in **Table 1**. There were no significant differences in age, sex, rs6971 polymorphism, disease severity, and disease duration between CBS and PSP patients. Both 4R tauopathy patient groups were matched for age, sex and rs6971 polymorphism when compared to controls. MoCA was significantly reduced in CBS (22.4±5.7, *p*=0.001) and PSP (22.2±4.7, *p*=0.004) patients when compared to controls (29.1±1.0).

**Table 1.** Demographics at the group level: 4RT = 4-repeat tauopathies; CBS = corticobasal syndrome; PSP = progressive supranuclear palsy; RS = Richardson syndrome; PSPRS = PSP rating scale; m = months; n = sample size; MoCA = Montreal Cognitive Assessment; SEADL = Schwab and England Activities of Daily Living; n.a. = not available; HAB = high affinity binder; MAB = medium affinity binder. Demographics were statistically tested by ANOVA or X^2^ test. * indicates p < 0.01

| Demographics | 4RT | CBS | PSP-RS | Controls |
| --- | --- | --- | --- | --- |
| n | 44 | 30 | 14 | 13 |
| Age | 68.3 ± 8.6 | 68.3 ± 9.1 | 68.8 ± 8.0 | 70.3 ± 7.5 |
| Sex | 24 ♀ / 20 ♂ | 16 ♀ / 14 ♂ | 8 ♀ / 6 ♂ | 7 ♀ / 6 ♂ |
| rs6971 | HAB: 30 / MAB: 14 | HAB: 22 / MAB: 8 | HAB: 8 / MAB: 6 | HAB: 5 / MAB: 8 |
| PSPRS | 31.8 ± 13.4 | 29.7 ± 13.4 | 36.5 ± 12.7 | n.a. |
| Disease duration (m) | 32.7 ± 26.0 | 30.8 ± 21.9 | 36.7 ± 33.8 | n.a. |
| MoCA | 22.4 ± 5.7* | 22.4 ± 4.7* | 22.2 ± 7.8 | 29.1 ± 1.0 |
| SEADL | 60.0 ± 18.2 | 60.7 ± 18.0 | 58.6 ± 19.2 | n.a. |
| Armstrong Criteria | - | 12 possible CBS, 18 probable CBS | - | - |
| MDS PSP Criteria | - | 7 not fulfilled, 4 suspected PSP-CBS, 19 possible PSP-CBS | 1 possible PSP, 13 probable PSP | - |

Predefined regions of interest indicated significantly higher TSPO-PET SUVr in most subcortical areas of CBS and PSP patients when compared to controls with the strongest differences in the globus pallidus internus (SUVr CBS: 1.039 [95%CI 1.000–1.078, *p*<0.001], SUVr PSP: 1.046 [95%CI: 0.990–1.101, *p*<0.001], SUVr controls: 0.861 [95%CI 0.802– 0.921]; **Table 2**). CBS patients also showed significantly elevated TSPO-PET SUVr versus controls in cortical regions of the frontal lobe, including the precentral gyrus, the middle frontal gyrus and the inferior frontal gyrus, whereas PSP patients did not show elevated TSPO-PET SUVr versus control subjects (**Table 2**).

**Table 2.**
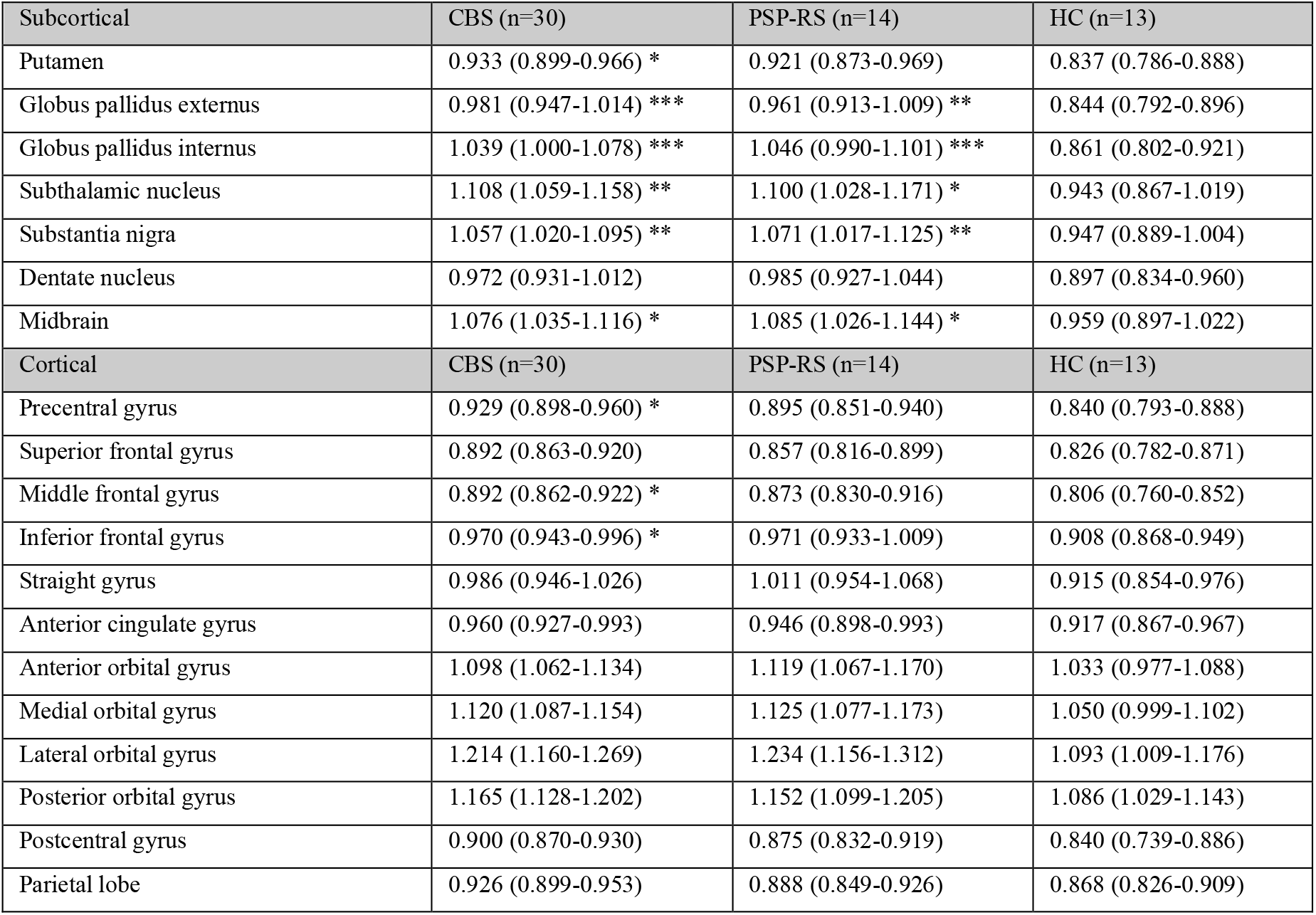
TSPO-PET quantification at the group level: Values represent regional group means of TSPO-PET SUVr (60-80 min time window) and their 95% confidence interval in predefined subcortical and cortical brain areas. Significance levels are indicated by *p<0.05, **p<0.01, ***p<0.001. P values were derived from multivariate analysis of variance with age, sex and rs6971 polymorphism as covariates and Bonferroni *post hoc* correction for multiple study groups. CBS = corticobasal syndrome; PSP = progressive supranuclear palsy; RS = Richardson syndrome; HC = controls without objectified memory impairment and with intact motor function

| Subcortical | CBS (n=30) | PSP-RS (n=14) | HC (n=13) |
| --- | --- | --- | --- |
| Putamen | 0.933 (0.899-0.966) * | 0.921 (0.873-0.969) | 0.837 (0.786-0.888) |
| Globus pallidus externus | 0.981 (0.947-1.014) *** | 0.961 (0.913-1.009) ** | 0.844 (0.792-0.896) |
| Globus pallidus internus | 1.039 (1.000-1.078) *** | 1.046 (0.990-1.101) *** | 0.861 (0.802-0.921) |
| Subthalamic nucleus | 1.108 (1.059-1.158) ** | 1.100 (1.028-1.171) * | 0.943 (0.867-1.019) |
| Substantia nigra | 1.057 (1.020-1.095) ** | 1.071 (1.017-1.125) ** | 0.947 (0.889-1.004) |
| Dentate nucleus | 0.972 (0.931-1.012) | 0.985 (0.927-1.044) | 0.897 (0.834-0.960) |
| Midbrain | 1.076 (1.035-1.116) * | 1.085 (1.026-1.144) * | 0.959 (0.897-1.022) |
| Cortical | CBS (n=30) | PSP-RS (n=14) | HC (n=13) |
| Precentral gyrus | 0.929 (0.898-0.960) * | 0.895 (0.851-0.940) | 0.840 (0.793-0.888) |
| Superior frontal gyrus | 0.892 (0.863-0.920) | 0.857 (0.816-0.899) | 0.826 (0.782-0.871) |
| Middle frontal gyrus | 0.892 (0.862-0.922) * | 0.873 (0.830-0.916) | 0.806 (0.760-0.852) |
| Inferior frontal gyrus | 0.970 (0.943-0.996) * | 0.971 (0.933-1.009) | 0.908 (0.868-0.949) |
| Straight gyrus | 0.986 (0.946-1.026) | 1.011 (0.954-1.068) | 0.915 (0.854-0.976) |
| Anterior cingulate gyrus | 0.960 (0.927-0.993) | 0.946 (0.898-0.993) | 0.917 (0.867-0.967) |
| Anterior orbital gyrus | 1.098 (1.062-1.134) | 1.119 (1.067-1.170) | 1.033 (0.977-1.088) |
| Medial orbital gyrus | 1.120 (1.087-1.154) | 1.125 (1.077-1.173) | 1.050 (0.999-1.102) |
| Lateral orbital gyrus | 1.214 (1.160-1.269) | 1.234 (1.156-1.312) | 1.093 (1.009-1.176) |
| Posterior orbital gyrus | 1.165 (1.128-1.202) | 1.152 (1.099-1.205) | 1.086 (1.029-1.143) |
| Postcentral gyrus | 0.900 (0.870-0.930) | 0.875 (0.832-0.919) | 0.840 (0.739-0.886) |
| Parietal lobe | 0.926 (0.899-0.953) | 0.888 (0.849-0.926) | 0.868 (0.826-0.909) |

Statistical parametric mapping validated higher TSPO labeling in the entire group of 4R tauopathy patients vs. control subjects, most pronounced in the midbrain and the pons, the globus pallidus, the dentate nucleus and motor and supplemental motor cortex (**Fig. 3A, Supplemental Table 1A**). Clusters in the basal ganglia, the midbrain, the precentral gyrus, the anterior cingulate gyrus, the inferior frontal gyrus, and the insula survived correction for multiple comparisons (**Supplemental Table 1A**). CBS patients showed stronger elevations of TSPO labeling in motor and supplemental motor cortices when compared to PSP, whereas subcortical elevations of TSPO labeling were of similar magnitude between CBS and PSP subgroups (compare **Fig. 3B** and **Fig. 3C, Supplemental Tables 1B,C**).

**Figure 3.**
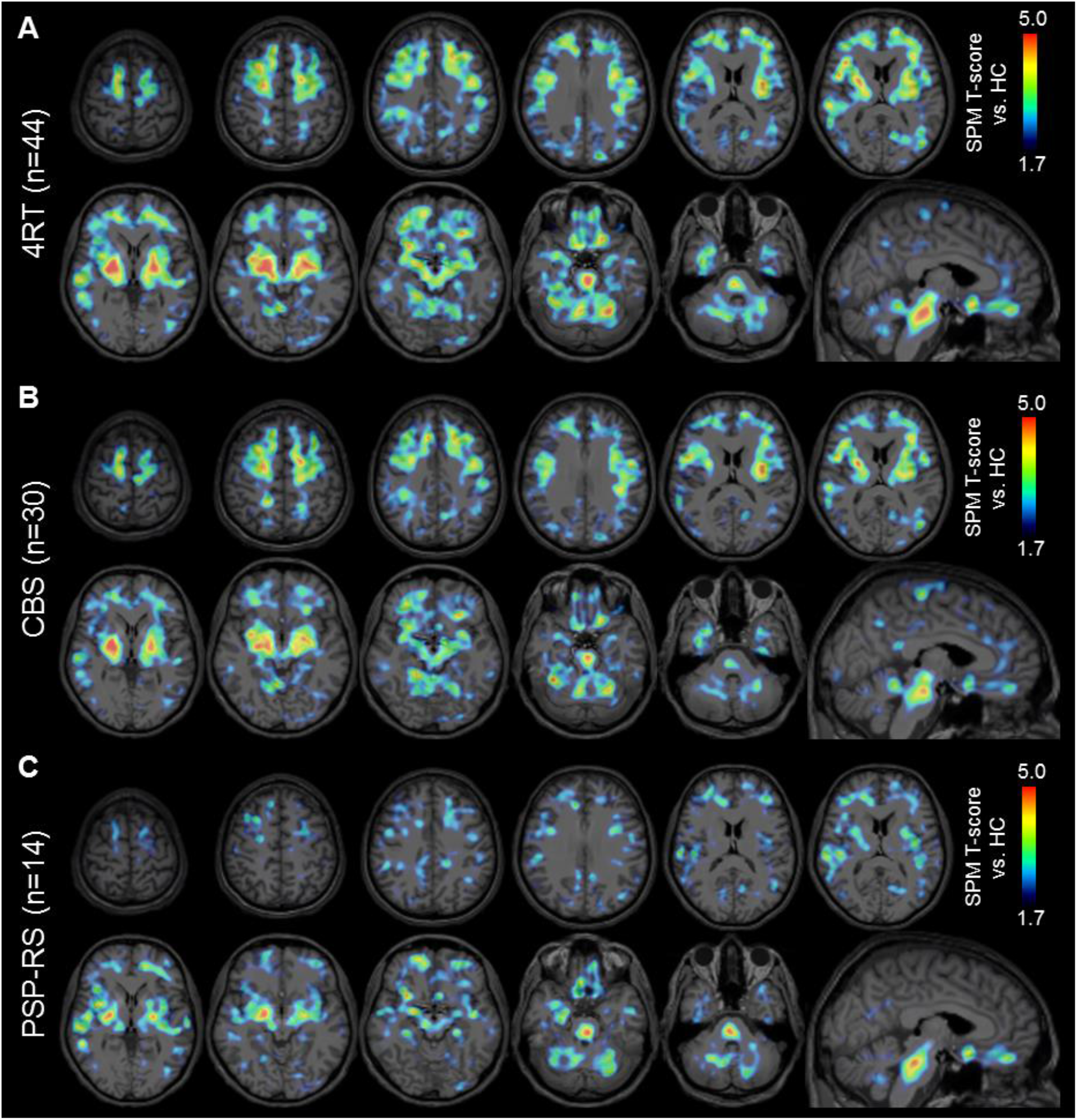
Voxel-based differences of TSPO-PET binding in 4R tauopathy patients: Color coding shows brain areas with increased TSPO-PET binding in groups of all 4R tauopathy patients (**A**, 4RT, n=44), corticobasal syndrome patients (**B**, CBS, n=30), and progressive supranuclear palsy Richardson syndrome patients (**C**, PSP-RS, n=14) when compared to controls (HC, n=13). T-statistics derive from an unpaired Student’s t-test with age, sex and rs6971 polymorphism as covariates as calculated by statistical parametric mapping (SPM). The lower threshold reflects a significance level of p=0.05, uncorrected for multiple comparisons. Extracerebral voxels are masked. T-score maps are projected in axial and sagittal planes upon a standard MRI atlas.

### TSPO labeling in 4R tauopathy patients allows identification in early disease stages

TSPO labeling in predefined regions of interest showed only weak to moderate associations with parameters of disease progression (**Table 3**). In CBS patients, significant partial correlations were observed for TSPO labeling in the substantia nigra with the PSPRS (*R*=- 0.463, *p*=0.015) and for TSPO labeling in the anterior cingulate cortex with the PSPRS (*R*=- 0.530, *p*=0.004) and the MoCA score (*R*=0.556, *p*=0.003). There were no significant associations of regional TSPO labeling and parameters of disease progression in PSP. Eighteen out of 30 patients with CBS indicated a right peripheral predominance of clinical symptoms, eleven showed a predominant left phenotype and one was affected equilaterally. Asymmetry indices of TSPO-PET in subcortical regions showed a significant lateralization to the hemisphere contralateral of the clinical phenotype (49/77, χ^2^=5.11, *p*=0.024) with highest agreement in the internal (15/18, χ^2^=8.10, *p*=0.004) and external (9/9, χ^2^=9.00, *p*=0.003) part of the globus pallidus. Asymmetry indices of TSPO-PET in the frontal cortex were not significantly associated with lateralization of the clinical phenotype 49/98, χ^2^=1.98, *p*=0.160). Given the weak associations of TSPO labeling with parameters of disease progression in contrast to large differences between 4R tauopathy patients and controls at the group level, we hypothesized a potential of TSPO-PET as a diagnostic biomarker in early disease stages. A multiregion classifier using a SUVr threshold >mean value ±2 standard deviations in controls indicated a sensitivity of 80% for the full cohort of CBS patients. Here, one positive target region exceeding the control threshold classified the subject positive for presence of a CBS (**Fig. 4A**). Importantly, the sensitivity in the sub-cohort of twelve patients with possible CBS was still 75%. This was also reflected by a similar magnitude of summed z-score vectors of TSPO labeling in target regions between possible and probable CBS (**Fig. 4B**). In line with single brain regions, there was no significant association between summed z-score vectors of TSPO labeling in target regions of CBS patients and parameters of disease progression (**Fig. 4C-E**). Sensitivity for PSP was 79% and there was only one outlier in the group of controls, indicating an overall specificity of 92% (**Fig. 4A**).

**Table 3.** Associations of TSPO-PET with parameters of disease progression in 4R tauopathies: Values represent partial correlation coefficients of TSPO-PET SUVr (60-80 min time window) with PSP Rating Scale, disease duration and MoCA. Significant associations are indicated by *p<0.05. P values were derived from partial correlation with age, sex and rs6971 polymorphism as covariates. CBS = corticobasal syndrome; PSP = progressive supranuclear palsy; RS = Richardson syndrome

|  | CBS (n=30) |  |  | PSP-RS (n=14) |  |  |
| --- | --- | --- | --- | --- | --- | --- |
|  | PSP<br>Rating<br>Scale | Disease<br>Duration | MoCA | PSP<br>Rating<br>Scale | Disease<br>Duration | MoCA |
| Subcortical |  |  |  |  |  |  |
| Putamen | -0.347 | 0.171 | 0.370 | 0.064 | -0.131 | 0.469 |
| Globus pallidus externus | -0.305 | 0.119 | 0.378 | 0.173 | 0.105 | 0.323 |
| Globus pallidus internus | -0.106 | 0.137 | 0.253 | 0.321 | -0.087 | 0.199 |
| Subthalamic nucleus | -0.203 | 0.149 | 0.146 | 0.406 | -0.048 | 0.055 |
| Substantia nigra | -0.463* | -0.036 | 0.287 | 0.243 | -0.094 | 0.087 |
| Dentate nucleus | -0.320 | -0.109 | 0.299 | 0.065 | 0.010 | 0.087 |
| Midbrain | -0.349 | 0.023 | 0.313 | 0.314 | -0.369 | 0.130 |
| Cortical |  |  |  |  |  |  |
| Precentral gyrus | 0.051 | 0.291 | 0.166 | 0.090 | -0.138 | 0.085 |
| Superior frontal gyrus | -0.016 | 0.117 | 0.054 | 0.031 | 0.451 | -0.063 |
| Middle frontal gyrus | 0.041 | 0.314 | 0.211 | 0.213 | 0.451 | -0.030 |
| Inferior frontal gyrus | -0.220 | 0.280 | 0.340 | 0.029 | 0.099 | 0.218 |
| Straight gyrus | -0.172 | -0.030 | 0.200 | 0.149 | 0.457 | -0.019 |
| Anterior cingulate gyrus | -0.530* | 0.052 | 0.556* | 0.024 | -0.086 | 0.246 |
| Anterior orbital gyrus | -0.360 | -0.068 | 0.251 | 0.353 | -0.044 | 0.055 |
| Medial orbital gyrus | -0.256 | -0.047 | 0.156 | 0.407 | 0.381 | 0.046 |
| Lateral orbital gyrus | -0.244 | 0.112 | 0.019 | 0.340 | -0.076 | -0.034 |
| Posterior orbital gyrus | -0.348 | 0.013 | 0.074 | 0.479 | -0.289 | 0.084 |
| Postcentral gyrus | 0.029 | 0.317 | 0.226 | 0.062 | -0.358 | 0.100 |
| Parietal lobe | -0.176 | 0.326 | 0.313 | 0.165 | -0.023 | 0.279 |

**Figure 4.**
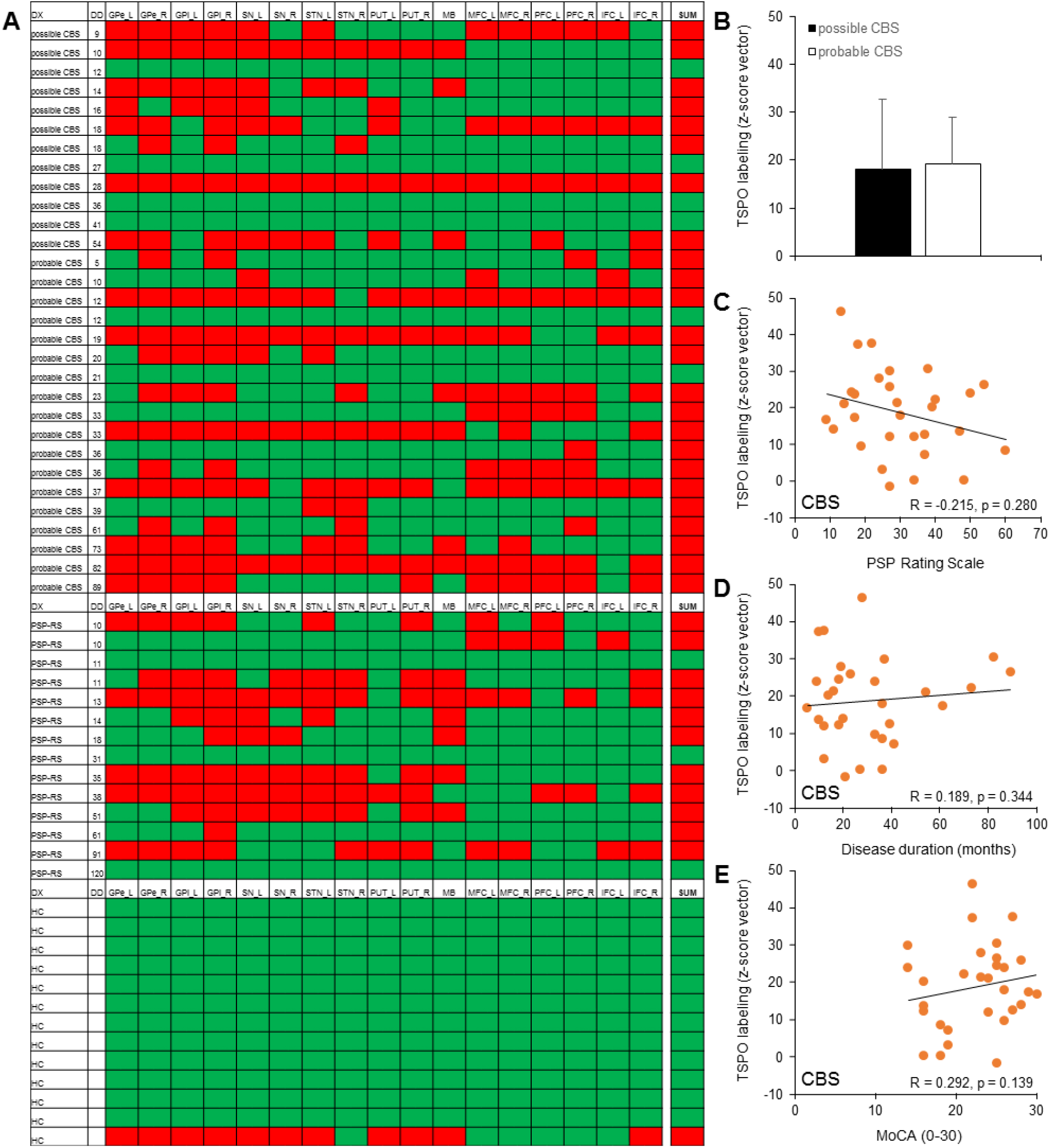
Multiregion classifier for diagnosis of 4R tauopathies by TSPO-PET. (**A**) Semi-quantitative classification (red = positive, green = negative) of 4R tauopathy target regions was performed by applying a mean value (MV) + 2 standard deviations (SD) threshold as obtained from the controls without objectified memory impairment and with intact motor function (HC) data. One single region defined the scan as global positive (SUM). Patients with corticobasal syndrome (CBS) and progressive supranuclear palsy Richardson syndrome (PSP-RS) are sorted according to possible or probable diagnosis and disease duration. (**B**) Comparison of z-score vectors (sum of regional z-score in regions with significant differences between 4R tauopathies and HC, see Table 2) of elevated TSPO labeling between possible and probable corticobasal syndrome diagnoses. (**C-E**) Associations between parameters of disease progression and multiregional TSPO labeling in corticobasal syndrome patients. R/p values derive from partial correlation, controlled for age, sex and rs6971 polymorphism. DX = clinical diagnosis; DD = disease duration; GPe = globus pallidus externus; GPi = globus pallidus internus; PUT = putamen; STN = subthalamic nucleus; SN = substantia nigra; MB = midbrain; DN = dentate nucleus; MFC = middle frontal cortex; PFC = prefrontal cortex; IFC = inferior frontal cortex; L = left; R = right; MoCA = Montreal Cognitive Assessment

### sTREM2 peaks at low TSPO labeling in CBS

A subpopulation of 18 patients with CBS and 12 controls was eligible for CSF sTREM2 measures. There was no difference in absolute sTREM2 levels between patients with CBS and controls (0.96 [95%CI 0.89–1.04] vs. 0.96 [95%CI 0.86–1.05], p=0.950, **Fig. 5A**). Measures of sTREM2 in patients with CBS as a function of TSPO labeling described an inverted U-shape with a peak at low TSPO-PET signal in 4R tauopathy target regions (**Fig. 5B** and **Fig. 5C**). In TSPO-positive patients (>2 SD of controls), there was a negative linear association between TSPO labeling and sTREM2 (R=-0.616, p=0.016). There was no significant association between parameters of disease progression and sTREM2 (**Fig. 5D-F**).

**Figure 5.**
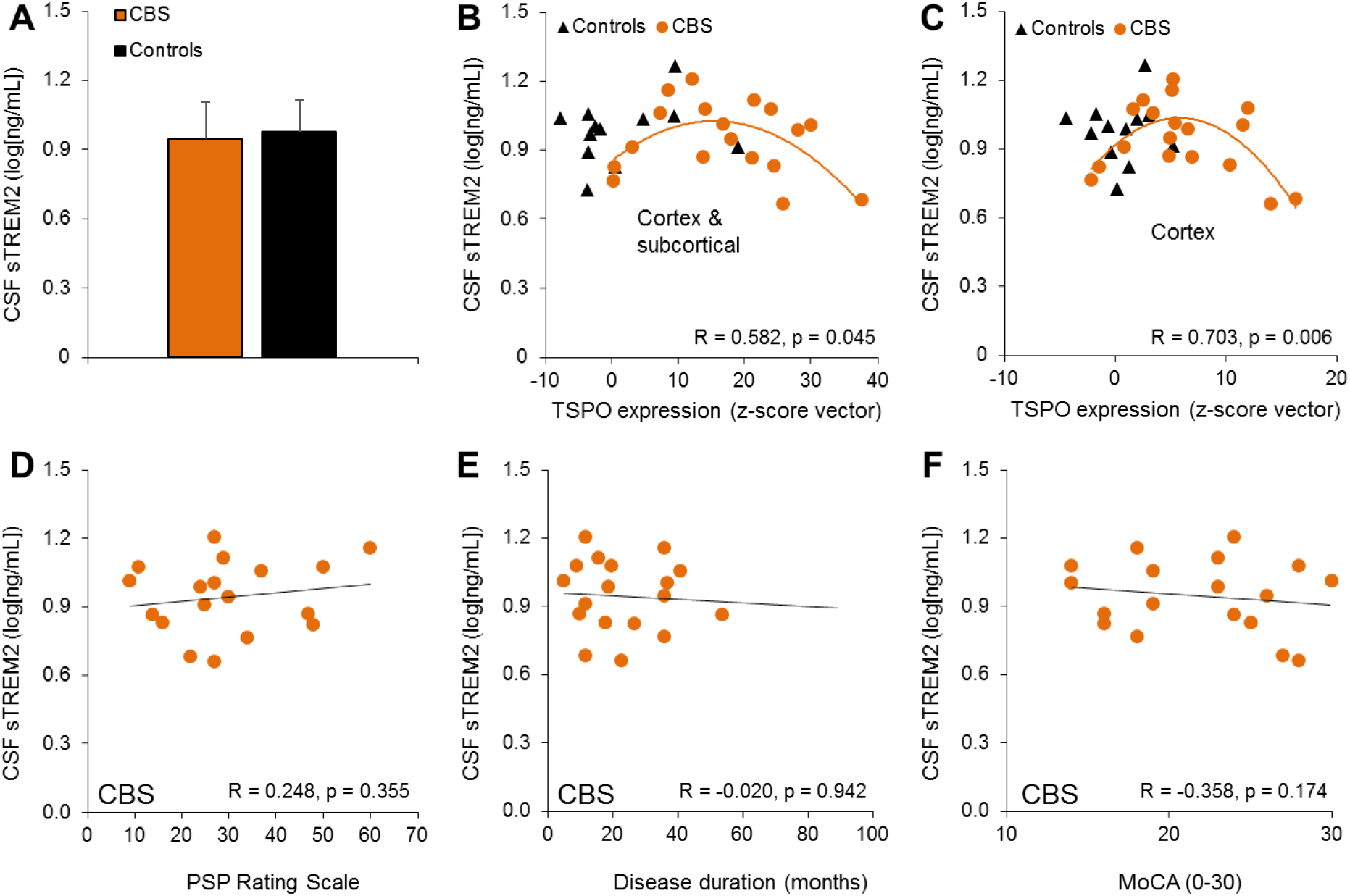
sTREM2 in patients with corticobasal syndrome. (**A**) Comparison of sTREM2 between patients with corticobasal syndrome (CBS) and controls, controlled for age and sex. (**B-C**) Associations between multiregional TSPO labeling and sTREM2 in corticobasal syndrome patients for all brain regions with significant differences in TSPO-PET (**B**) and a subanalysis in cortical regions (**C**). R/p values derive from quadratic fits. (**D-F**) Associations between parameters of disease progression and sTREM2 in corticobasal syndrome patients. R/p values derive from partial correlation, controlled for age and sex. MoCA = Montreal Cognitive Assessment. CSF = cerebrospinal fluid

## Discussion

We provide the first human study using a fluorinated TSPO-PET radiotracer in a meaningful sized cohort of CBS and PSP patients. Our translational data in a 4R tauopathy mouse model verified that ^18^F-GE-180 detects 4R tau related microglial activation with high specificity. Dynamic imaging and methodological workup in humans indicated feasibility of TSPO-PET imaging with low patient effort and simplified quantification approaches. CBS and PSP patients revealed elevated TSPO labeling in subcortical brain regions, whereas CBS patients additionally showed stronger elevation of TSPO labeling in the forebrain. Cross-sectional correlation analyses indicated only minor associations of TSPO labeling with parameters of disease progression but a potential of TSPO-PET as an early disease stage biomarker in CBS.

To the best of our knowledge there are only two published reports on TSPO-PET imaging in 4R tauopathies, both only including PSP patients^14, 15^, whereas our study is the first including CBS. A study from 2006 investigated a small cohort of four PSP using ^11^C-PK11195, implementing serial imaging in two patients^14^. Their baseline findings of the strongest TSPO-PET increases in basal ganglia and brainstem are in line with our observations in 14 PSP patients, whereas we could not confirm differences in the frontal cortex. A more recent study scanned 16 PSP patients with a mean PSPRS score of 40.8 with ^11^C-PK11195 together with an equal number of Alzheimer’s disease patients and 13 controls and found higher TSPO labeling in putamen, thalamus and pallidum in PSP compared to controls, but no binding differences vs. Alzheimer’s disease^15^. Our regional findings in 14 PSP patients of similar disease severity (mean PSPRS score: 36.5) confirm the earlier observations using a third generation TSPO tracer. Our novel TSPO-PET data in CBS mirror the subcortical findings of PSP, but reveal an additional elevation of TSPO labeling in the neocortex with the strongest signal in the motor and supplemental motor areas of the forebrain. This fits well to the pronounced cortical phenotype of CBS compared to PSP and is also in line with the concept of overlapping neuropathology among different 4R tauopathies^4^.

Recently, ^18^F-GE-180 to assess TSPO labeling in patients with glioma and multiple sclerosis was criticized because of inferior image quality and limited uptake across the blood brain barrier^33-35^. Based on our preclinical experience, we were aware of a high correlation of the ^18^F-GE-180 PET signal with immunohistochemistry in mouse models of neurodegenerative disorders despite the low brain penetration of the tracer^13, 32, 36^. Although we deemed it unlikely that a parallel blood brain barrier phenomenon caused these strong associations, we made use of pharmacological microglial depletion to prove specificity of the tracer for activated microglia in presence of 4R tau. As expected, we observed a clear decrease of the ^18^F-GE-180 PET signal in microglia depleted P301S mice when compared to their own individual baseline. This strongly supports the claim of a specific PET signal *in vivo*. The translational application of ^18^F-GE-180 in 4R tauopathy patients consecutively revealed a pattern of TSPO labeling fitting to the neuropathological topologies of CBS and PSP in former autopsy studies^37-39^.

Even without correction for multiple comparisons, we found only a few weak associations between measures of disease progression and regional TSPO labeling in CBS and no associations in PSP, controlling for age, sex and rs6971 polymorphism. Yet, our ^18^F-GE-180 PSP findings were roughly in line with positive associations of ^11^C-PK11195 binding and disease severity in a cohort of 16 PSP patients, showing a similarly large inter-individual heterogeneity of TSPO labeling at a given PSPRS score^15^. In contrast, the regional associations of TSPO labeling with parameters of disease progression in CBS showed a higher TSPO-PET signal in early stages of disease progression. We found a sensitivity of 75% in possible CBS patients and even higher sensitivity for probable CBS and PSP vs. controls using a simple semi-quantitative multiregion classifier, underlining the potential of ^18^F-GE-180 TSPO-PET as a supportive biomarker for diagnosis of 4R tauopathies and improving limited sensitivity of clinical diagnosis^40^. Despite the lacking association between TSPO labeling and parameters of disease progression, we find a clear agreement between asymmetries of subcortical TSPO labeling and contralateral clinical predominance of the CBS phenotype. This supports the important role of microglial activation in the pathophysiological cascade of 4R tauopathies^6^. A subpopulation of patients with CBS and controls was eligible for CSF sTREM2 measures and our limited data do not support sTREM2 as a diagnostic biomarker in CBS since there was no difference between patients and controls. However, we find an interesting negative association between TSPO-PET and sTREM2 in patients with CBS after a sTREM2 peak at low levels of TSPO. Speculatively, this could indicate a functional burnout of microglia with increasing activity and deserves further longitudinal exploration. The observed cross-sectional heterogeneity of TSPO labeling in 4R tauopathy patients will enable predictive analyses, aiming to elucidate if high or low TSPO labeling at baseline is associated with better clinical outcome. Taken together, longitudinal PET studies and predictive analyses of the clinical course are needed for further exploration of TSPO-PET as a biomarker in immunomodulatory trials. The current 4R tauopathy cohort will be followed clinically and by serial TSPO-PET to address this question.

Among the limitations of our study we note missing autopsy validation of the studied clinically diagnosed 4R tauopathy cases. However, the high specificity of a clinical 4R tauopathy diagnosis predicts a low number of false positive cases in the studied dataset^40^. We acknowledge as another limitation that we did not include arterial sampling in the dynamic imaging protocol, thus, although we deem it unlikely, we cannot exclude effects from differences in plasma fractions and tracer metabolism among our study groups. β-amyloid PET was not performed in the PSP arm of the study due to the high clinical probability of predicting a 4R tauopathy. Thus, we cannot rule out frequent^41^ β-amyloid co-pathology and a subsequent impact on microglial activation in PSP patients, which however does not have a major impact on clinical progression.

To conclude, TSPO-PET imaging closely reflects the expected topology of microglial activation in 4R tauopathies and shows potential as a neuroinflammation biomarker. *In vivo* assessment of TSPO labeling has a potential to support early diagnosis of 4R tauopathies, facilitating increased sensitivity. Longitudinal studies are needed to explore the value of TSPO-PET imaging as a progression biomarker in 4R tauopathies.

## Data Availability

Data of this manuscript is available from the corresponding author upon reasonable request.

## Acknowledgements

We thank all of our patients, their care-givers, cyclotron, radiochemistry and the PET imaging crew. GE Healthcare made GE-180 cassettes available through an early-access model. The authors thank Plexxikon Inc. for providing PLX5622.

## Funding

This work was funded by the Deutsche Forschungsgemeinschaft (DFG, German Research Foundation) to P.B. and N.A. – project number 421887978, to C.W. – project number DFG WE2298/10-1, 422182557, and to A.R. and M.B. – project numbers BR4580/1-1/ RO5194/1-1. This project was also supported by the German Center for Neurodegenerative Diseases (DZNE, DescribePSP Study), the German Parkinson’s Association (DPG, ProPSP Study) and the Hirnliga e.V. (Manfred-Strohscheer-Stiftung). P.B., G.U.H., C.H., J.H. and R.P. were supported by the Deutsche Forschungsgemeinschaft (DFG, German Research Foundation) under Germany’s Excellence Strategy within the framework of the Munich Cluster for Systems Neurology (EXC 2145 SyNergy – ID 390857198). G.U.H. was also funded by the NOMIS foundation (FTLD project). The Lüneburg Heritage has supported the work of C.P.

## Competing Interests

M.B. received speaker honoraria from GE healthcare and LMI and is an advisor of LMI. G.U.H. received research support from GE Healthcare and Neuropore; has ongoing research collaborations with Orion and Prothena; serves as a consultant for AbbVie, AlzProtect, Asceneuron, Biogen, Biohaven, Lundbeck, Novartis, Roche, Sanofi, UCB; received honoraria for scientific presentations from AbbVie, Biogen, Roche, Teva, UCB, and Zambon; and holds a patent on PERK Activation for the Treatment of Neurodegenerative Diseases (PCT/EP2015/068734). C.H. is chief scientific advisor of ISAR biosciences and collaborates with DENALI therapeutics. R.P. is on the advisory board for Biogen, has consulted for Eli Lilly, is a grant recipient from Janssen Pharmaceutica and Boehringer Ingelheim, and has received speaker honoraria from Janssen-Cilag, Pfizer and Biogen. J.L. reports speaker fees from Bayer Vital, consulting fees from Axon Neuroscience, author fees from Thieme medical publishers and W. Kohlhammer GmbH medical publishers, non-financial support from Abbvie and compensation for duty as part-time CMO from MODAG GmbH, all outside the submitted work. All other authors do not report a conflict of interest.

## Author contributions

C.P., S.S., A.R., P.B., J.H., R.P, C.H., J.L., G.U.H., and M.B. conceived the study and analyzed the results. C.P., J.S., J.L., G.U.H. and M.B. wrote the manuscript with help from C.H. and further input from all co-authors. J.S., L.B., S.H., J.S., A.F., and A.N. performed human PET scans and their analysis. E.W., C.P., K.B., A.D., B.R., R.P, J.L. and G.H. recruited patients, examined patients and controls and analyzed clinical data. F.R., F.E., G.B., T.B. and Y.S. performed small animal PET experiments and small animal immunohistochemistry. E.M. and C.H. performed sTREM2 measures and analysis as well as the interpretation of sTREM2 results in conjunction with TSPO-PET. M.U., N.L.A., C.W. and R.R. performed TSPO polymorphism genotyping and analysis. J.S., L.B., A.R., P.B. and M.B. interpreted human PET data.

## Supplementary Material

**Supplementary Table 1.** Findings of voxel-wise ^18^F-GE-180 PET analyses for the contrast of 4-repeat tauopathy patients (4RT), patients with corticobasal syndrome (CBS) and patients with progressive supranuclear palsy Richardson syndrome (PSP-RS) against controls. Statistical parametric mapping (SPM) t-statistics derive from an unpaired Student’s t-test with age, sex and rs6971 polymorphism as covariates. k > 100 voxels. Data indicated in bold font indicate a cluster and non-bold data identify sub-peaks within the same cluster. Regions highlighted in grey survived strict family-wise correction for multiple testing (FWE). Locations are reported as the regions in a cube range within ±1 mm. Numeric data are Talairach and Tournoux coordinates transformed from Montreal Neurology Institute space (x, y, z; mm). R = right. L = left. BA = Brodmann. uncorr. = uncorrected for multiple comparisons.

| 4RT: Group comparison vs. controls | Talairach and |  |  |  |  |  |
| --- | --- | --- | --- | --- | --- | --- |
|  | Tournoux coordinates |  |  |  |  |  |
| Location (cube range ± 1 mm) | x | y | z | peak | p value | p-value |
|  |  |  |  | t-value | uncorr. | FWE |
| R Lentiform Nucleus<br>R Globus Pallidus<br>R Putamen | 23 | -9 | 3 | 6.28 | 0.000 | 0.002 |
| R Anterior Cingulate Gyrus | 16 | 15 | 39 | 5.73 | 0.000 | 0.011 |
| L Lentiform Nucleus<br>L Globus Pallidus | -21 | -10 | 0 | 5.71 | 0.000 | 0.012 |
| R Insula<br>R Precentral Gyrus<br>R Inferior Frontal Gyrus | 42 | 18 | 9 | 5.46 | 0.000 | 0.026 |
| L Brainstem, Midbrain<br>R Brainstem, Midbrain | 0 | -25 | -14 | 5.45 | 0.000 | 0.027 |
| L Inferior Frontal Gyrus, BA 47 | -18 | 17 | -17 | 5.15 | 0.000 | 0.064 |
| L Cerebellum, Declive | -25 | -62 | -16 | 5.04 | 0.000 | 0.090 |
| R Claustrum<br>R Insula | 36 | -3 | -1 | 5.02 | 0.000 | 0.094 |
| L Cerebellum, Declive | -21 | -64 | -14 | 4.99 | 0.000 | 0.102 |
| R Lentiform Nucleus<br>R Putamen | 27 | -2 | 10 | 4.98 | 0.000 | 0.105 |
| L Anterior Cingulate Gyrus, BA 32 | -13 | 11 | -37 | 4.93 | 0.000 | 0.122 |
| R Medial Frontal Gyrus, BA 6, BA 8 | 10 | 29 | 37 | 4.90 | 0.000 | 0.131 |
| L Inferior Frontal Gyrus, BA 47<br>L Middle Frontal Gyrus, BA 11 | -26 | 23 | -14 | 4.89 | 0.000 | 0.135 |

**Supplementary Table 2.**
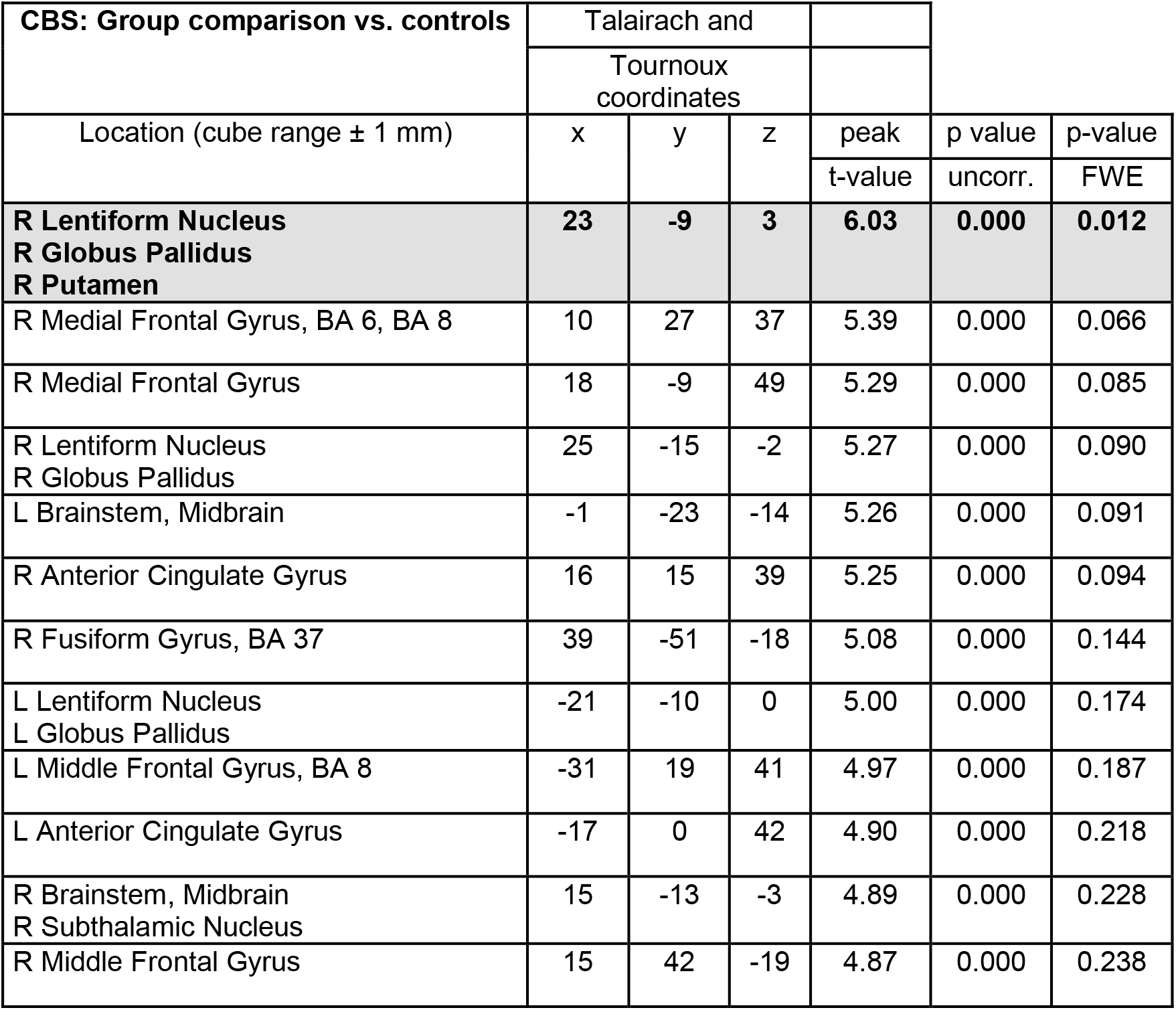
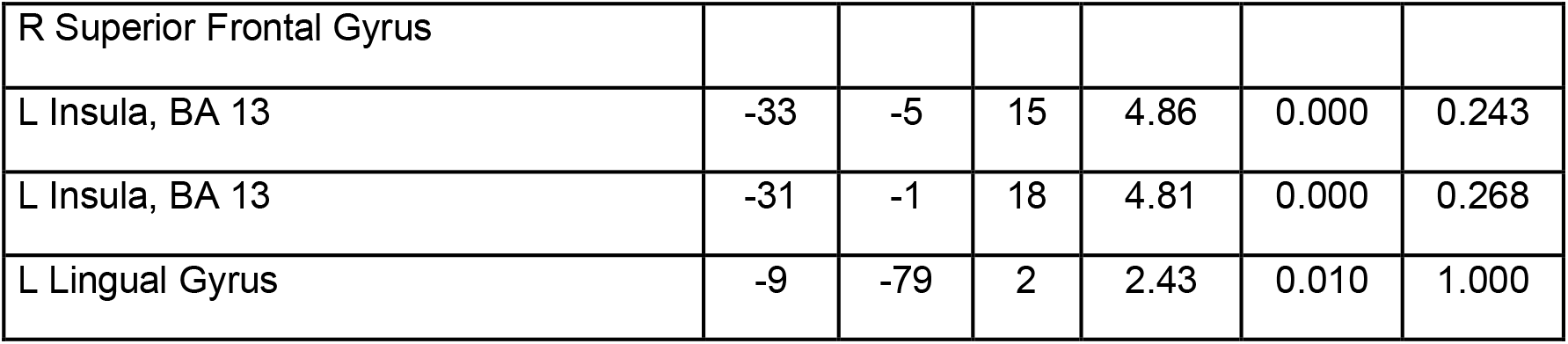
Findings of voxel-wise ^18^F-GE-180 PET analyses for the contrast of 4-repeat tauopathy patients (4RT), patients with corticobasal syndrome (CBS) and patients with progressive supranuclear palsy Richardson syndrome (PSP-RS) against controls. Statistical parametric mapping (SPM) t-statistics derive from an unpaired Student’s t-test with age, sex and rs6971 polymorphism as covariates. k > 100 voxels. Data indicated in bold font indicate a cluster and non-bold data identify sub-peaks within the same cluster. Regions highlighted in grey survived strict family-wise correction for multiple testing (FWE). Locations are reported as the regions in a cube range within ±1 mm. Numeric data are Talairach and Tournoux coordinates transformed from Montreal Neurology Institute space (x, y, z; mm). R = right. L = left. BA = Brodmann. uncorr. = uncorrected for multiple comparisons.

**Supplementary Table 3.**
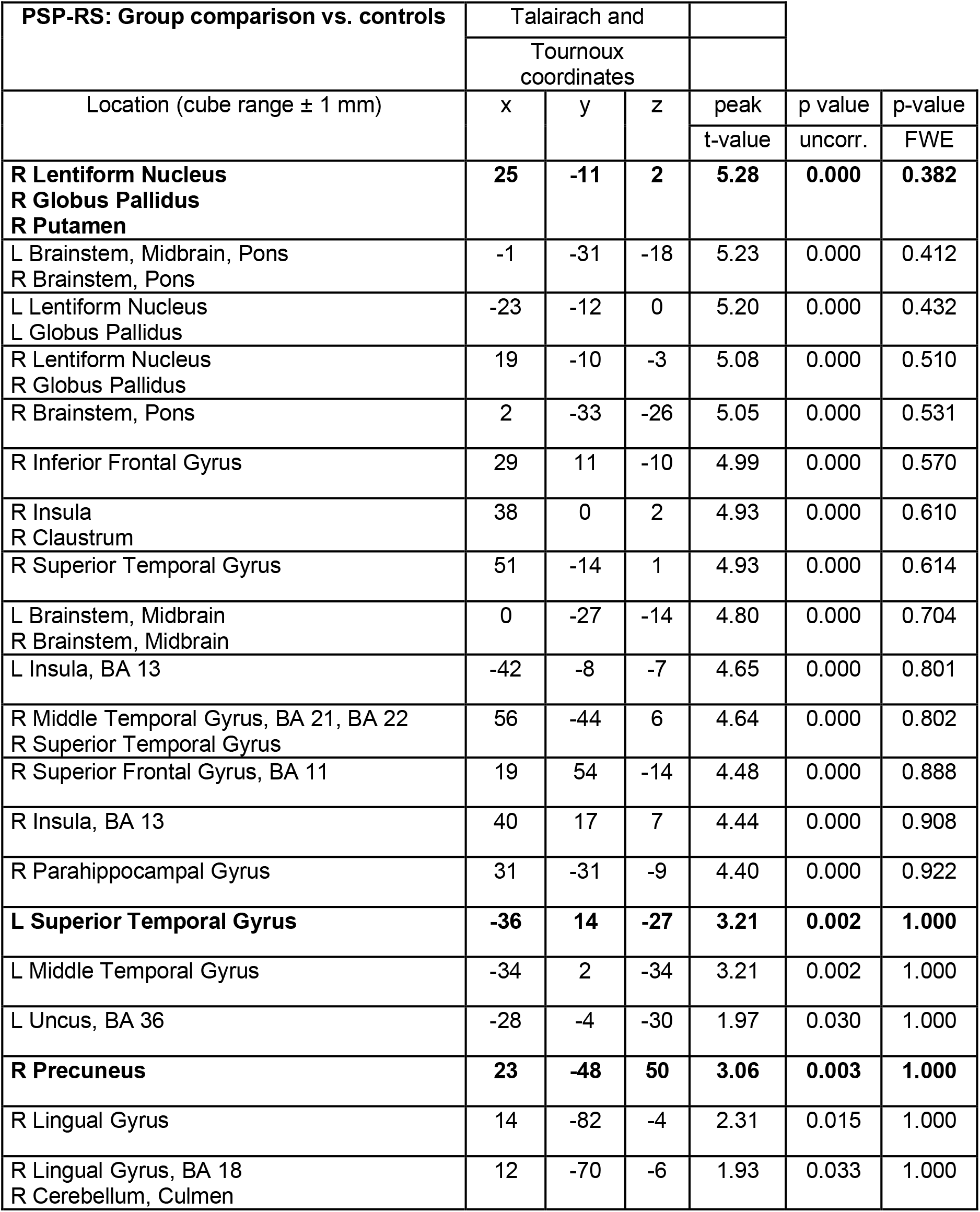
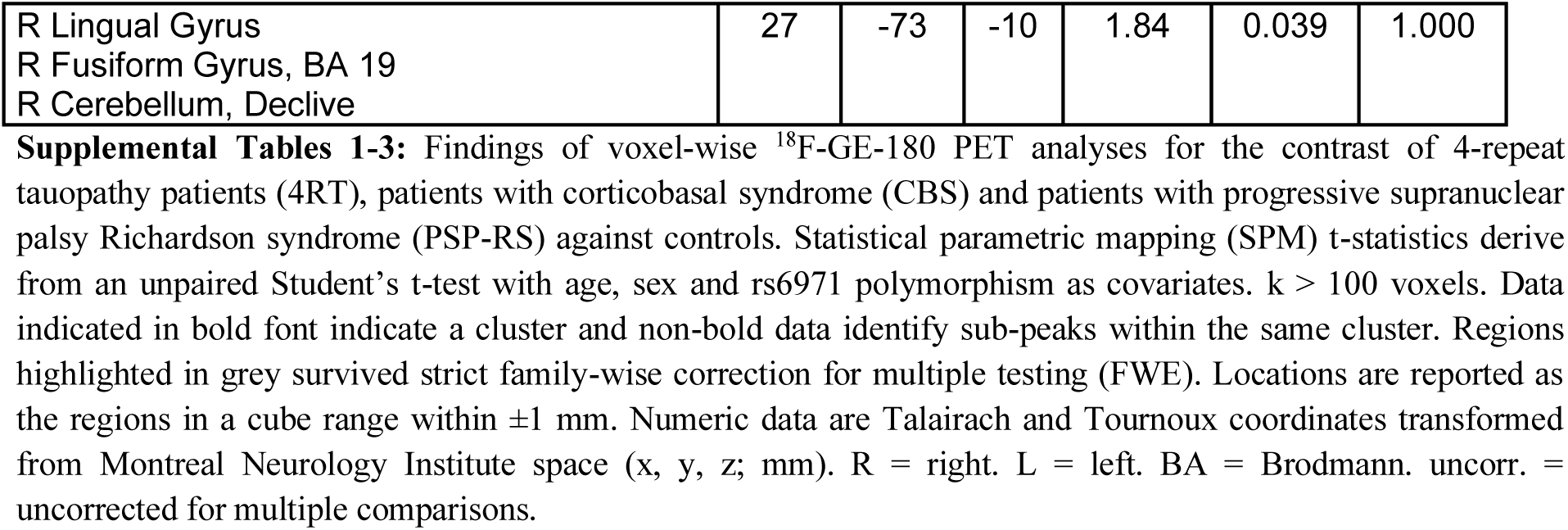
Findings of voxel-wise ^18^F-GE-180 PET analyses for the contrast of 4-repeat tauopathy patients (4RT), patients with corticobasal syndrome (CBS) and patients with progressive supranuclear palsy Richardson syndrome (PSP-RS) against controls. Statistical parametric mapping (SPM) t-statistics derive from an unpaired Student’s t-test with age, sex and rs6971 polymorphism as covariates. k > 100 voxels. Data indicated in bold font indicate a cluster and non-bold data identify sub-peaks within the same cluster. Regions highlighted in grey survived strict family-wise correction for multiple testing (FWE). Locations are reported as the regions in a cube range within ±1 mm. Numeric data are Talairach and Tournoux coordinates transformed from Montreal Neurology Institute space (x, y, z; mm). R = right. L = left. BA = Brodmann. uncorr. = uncorrected for multiple comparisons.

## Notes

### Author Declarations

Human PET data analyses (ethics-applications: 17-569 & 19-022) were approved by the local institutional ethics committee (Ethics committee of the medical faculty, Ludwig-Maximilians-University, Munich, Germany). All participants provided written informed consent according to the Declaration of Helsinki. All animal experiments were performed in compliance with the National Guidelines for Animal Protection, Germany and with the approval of the regional animal committee (Regierung von Oberbayern) overseen by a veterinarian.

## References

1. Rosler TW, Tayaranian Marvian A, Brendel M, et al. Four-repeat tauopathies. Prog Neurobiol. 2019 Sep;180:101644.

2. Armstrong MJ, Litvan I, Lang AE, et al. Criteria for the diagnosis of corticobasal degeneration. Neurology. 2013 Jan 29;80(5):496–503.

3. Steele JC, Richardson JC, Olszewski J. Progressive Supranuclear Palsy. A Heterogeneous Degeneration Involving the Brain Stem, Basal Ganglia and Cerebellum with Vertical Gaze and Pseudobulbar Palsy, Nuchal Dystonia and Dementia. Arch Neurol. 1964 Apr;10:333–59.

4. Höglinger GU. Is it useful to classify progressive supranuclear palsy and corticobasal degeneration as different disorders? No. Movement disorders clinical practice. 2018;5(2):141.

5. Lee SE, Rabinovici GD, Mayo MC, et al. Clinicopathological correlations in corticobasal degeneration. Annals of neurology. 2011 Aug;70(2):327–40.

6. Ferrer I, López-González I, Carmona M, et al. Glial and neuronal tau pathology in tauopathies: characterization of disease-specific phenotypes and tau pathology progression. Journal of Neuropathology & Experimental Neurology. 2014;73(1):81–97.

7. Martini-Stoica H, Cole AL, Swartzlander DB, et al. TFEB enhances astroglial uptake of extracellular tau species and reduces tau spreading. J Exp Med. 2018 Sep 3;215(9):2355–77.

8. Perea JR, Llorens-Martin M, Avila J, Bolos M. The Role of Microglia in the Spread of Tau: Relevance for Tauopathies. Front Cell Neurosci. 2018;12:172.

9. Asai H, Ikezu S, Tsunoda S, et al. Depletion of microglia and inhibition of exosome synthesis halt tau propagation. Nature neuroscience. 2015 Nov;18(11):1584–93.

10. Ising C, Venegas C, Zhang S, et al. NLRP3 inflammasome activation drives tau pathology. Nature. 2019 Nov;575(7784):669–73.

11. Stefaniak J, O’Brien J. Imaging of neuroinflammation in dementia: a review. J Neurol Neurosurg Psychiatry. 2016 Jan;87(1):21–8.

12. Liu B, Le KX, Park MA, et al. In Vivo Detection of Age- and Disease-Related Increases in Neuroinflammation by 18F-GE180 TSPO MicroPET Imaging in Wild-Type and Alzheimer’s Transgenic Mice. J Neurosci. 2015 Nov 25;35(47):15716–30.

13. Parhizkar S, Arzberger T, Brendel M, et al. Loss of TREM2 function increases amyloid seeding but reduces plaque-associated ApoE. Nature neuroscience. 2019 Feb;22(2):191–204.

14. Gerhard A, Trender-Gerhard I, Turkheimer F, Quinn NP, Bhatia KP, Brooks DJ. In vivo imaging of microglial activation with [11C](R)-PK11195 PET in progressive supranuclear palsy. Mov Disord. 2006 Jan;21(1):89–93.

15. Passamonti L, Rodriguez PV, Hong YT, et al. [(11)C]PK11195 binding in Alzheimer disease and progressive supranuclear palsy. Neurology. 2018 May 29;90(22):e1989–e96.

16. Allen B, Ingram E, Takao M, et al. Abundant tau filaments and nonapoptotic neurodegeneration in transgenic mice expressing human P301S tau protein. J Neurosci. 2002 Nov 1;22(21):9340–51.

17. Brendel M, Probst F, Jaworska A, et al. Glial Activation and Glucose Metabolism in a Transgenic Amyloid Mouse Model: A Triple Tracer PET Study. J Nucl Med. 2016 Feb 18.

18. Brendel M, Focke C, Blume T, et al. Time Courses of Cortical Glucose Metabolism and Microglial Activity Across the Life Span of Wild-Type Mice: A PET Study. J Nucl Med. 2017 Dec;58(12):1984–90.

19. Hoglinger GU, Respondek G, Stamelou M, et al. Clinical diagnosis of progressive supranuclear palsy: The movement disorder society criteria. Mov Disord. 2017 Jun;32(6):853–64.

20. Respondek G, Klockgether T, Spottke A, Höglinger G. German prospective studies on PSP: ProPSP and DESCRIBE-PSP. Poster presented at Deutscher Kongress für Parkinson und Bewegungsstörungen, Düsseldorf. https://d-nb.info/1181570069/34. 2019.

21. Logan J. A review of graphical methods for tracer studies and strategies to reduce bias.Nucl Med Biol. 2003 Nov;30(8):833–44.

22. Vomacka L, Albert NL, Lindner S, et al. TSPO imaging using the novel PET ligand [(18)F]GE-180: quantification approaches in patients with multiple sclerosis. EJNMMI Res. 2017 Oct 26;7(1):89.

23. Feeney C, Scott G, Raffel J, et al. Kinetic analysis of the translocator protein positron emission tomography ligand [18F]GE-180 in the human brain. Eur J Nucl Med Mol Imaging. 2016 Nov;43(12):2201–10.

24. Brendel M, Barthel H, Van Eimeren T, et al. 18F-PI2620 Tau-PET in progressive supranuclear palsy: A multi-center evaluation. JAMA Neurology. 2020:in press.

25. Keuken MC, Bazin PL, Backhouse K, et al. Effects of aging on T(1), T(2)*, and QSM MRI values in the subcortex. Brain Struct Funct. 2017 Aug;222(6):2487–505.

26. Brendel M, Schonecker S, Hoglinger G, et al. [(18)F]-THK5351 PET Correlates with Topology and Symptom Severity in Progressive Supranuclear Palsy. Front Aging Neurosci. 2017;9:440.

27. Hammers A, Allom R, Koepp MJ, et al. Three-dimensional maximum probability atlas of the human brain, with particular reference to the temporal lobe. Hum Brain Mapp. 2003 Aug;19(4):224–47.

28. Owen DR, Gunn RN, Rabiner EA, et al. Mixed-affinity binding in humans with 18-kDa translocator protein ligands. J Nucl Med. 2011 Jan;52(1):24–32.

29. Owen DR, Yeo AJ, Gunn RN, et al. An 18-kDa translocator protein (TSPO) polymorphism explains differences in binding affinity of the PET radioligand PBR28. J Cereb Blood Flow Metab. 2012 Jan;32(1):1–5.

30. Kleinberger G, Yamanishi Y, Suarez-Calvet M, et al. TREM2 mutations implicated in neurodegeneration impair cell surface transport and phagocytosis. Sci Transl Med. 2014 Jul 2;6(243):243ra86.

31. Suarez-Calvet M, Morenas-Rodriguez E, Kleinberger G, et al. Early increase of CSF sTREM2 in Alzheimer’s disease is associated with tau related-neurodegeneration but not with amyloid-beta pathology. Mol Neurodegener. 2019 Jan 10;14(1):1.

32. Brendel M, Probst F, Jaworska A, et al. Glial Activation and Glucose Metabolism in a Transgenic Amyloid Mouse Model: A Triple-Tracer PET Study. J Nucl Med. 2016 Jun;57(6):954–60.

33. Zanotti-Fregonara P, Pascual B, Rostomily RC, et al. Anatomy of (18)F-GE180, a failed radioligand for the TSPO protein. Eur J Nucl Med Mol Imaging. 2020 Feb 22.

34. Zanotti-Fregonara P, Veronese M, Rizzo G, Pascual B, Masdeu JC, Turkheimer FE. Letter to the Editor re: Confirmation of Specific Binding of the 18-kDa Translocator Protein (TSPO) Radioligand [(18)F]GE-180: a Blocking Study Using XBD173 in Multiple Sclerosis Normal Appearing White and Grey Matter. Mol Imaging Biol. 2020 Feb;22(1):10–2.

35. Zanotti-Fregonara P, Veronese M, Pascual B, Rostomily RC, Turkheimer F, Masdeu JC. The validity of (18)F-GE180 as a TSPO imaging agent. Eur J Nucl Med Mol Imaging. 2019 Jun;46(6):1205–7.

36. Kleinberger G, Brendel M, Mracsko E, et al. The FTD-like syndrome causing TREM2 T66M mutation impairs microglia function, brain perfusion, and glucose metabolism. EMBO J. 2017 Jul 3;36(13):1837–53.

37. Williams DR, Holton JL, Strand C, et al. Pathological tau burden and distribution distinguishes progressive supranuclear palsy-parkinsonism from Richardson’s syndrome. Brain. 2007 Jun;130(Pt 6):1566–76.

38. Williams DR, Lees AJ. Progressive supranuclear palsy: clinicopathological concepts and diagnostic challenges. The Lancet Neurology. 2009 Mar;8(3):270–9.

39. Dickson DW. Neuropathologic differentiation of progressive supranuclear palsy and corticobasal degeneration. J Neurol. 1999 Sep;246 Suppl 2:II6–15.

40. Respondek G, Grimm MJ, Piot I, et al. Validation of the movement disorder society criteria for the diagnosis of 4-repeat tauopathies. Mov Disord. 2020 Jan;35(1):171–6.

41. Jecmenica Lukic M, Kurz C, Respondek G, et al. Copathology in Progressive Supranuclear Palsy: Does It Matter? Mov Disord. 2020 Mar 3.

